# Variants of concern are overrepresented among post-vaccination breakthrough infections of SARS-CoV-2 in Washington State

**DOI:** 10.1101/2021.05.23.21257679

**Authors:** Abbye E. McEwen, Seth Cohen, Chloe Bryson-Cahn, Catherine Liu, Steven A. Pergam, John Lynch, Adrienne Schippers, Kathy Strand, Estella Whimbey, Nandita S Mani, Allison J. Zelikoff, Vanessa A. Makarewicz, Elizabeth R. Brown, Shah A. Mohamed Bakhash, Noah R. Baker, Jared Castor, Robert J. Livingston, Meei-Li Huang, Keith R. Jerome, Alexander L. Greninger, Pavitra Roychoudhury

## Abstract

Across 20 vaccine breakthrough cases detected at our institution, all 20 (100%) infections were due to variants of concern (VOC) and had a median Ct of 20.2 (IQR=17.1-23.3). When compared to 5174 contemporaneous samples sequenced in our laboratory, VOC were significantly enriched among breakthrough infections (p < .05).

## Main text

Vaccination against SARS-CoV-2 elicits an immune response capable of potently neutralizing the virus. Multiple SARS-CoV-2 vaccines have been approved for use in humans through emergency use authorization or are in phase 3 clinical trials. Most of these vaccines use a recombinant spike protein derived from the first sequenced (Wuhan) strain from January 2020. The most widely used vaccines (mRNA-based mRNA-1273 and BNT162b2) have shown up to 95% efficacy at preventing clinical cases and up to 100% efficacy in preventing severe disease [1]. However, there is growing concern that emerging variants may escape vaccine-induced immunity and result in breakthrough infections in fully vaccinated individuals [2].

Since emerging in humans, SARS-CoV-2 is now defined by a variety of lineages harboring distinctive genetic changes in the spike protein. Variants of concern (VOCs) are those strains that show evidence of increased transmissibility, more severe disease, reduced neutralization by antibodies elicited by past infection or vaccination, reduced efficacy of treatments, or failures in diagnostic detection. As of May 20, 2021, there are currently 4 VOC identified by the US SARS-CoV-2 Interagency Group (P.1, B.1.351, B.1.427, and B.1.429) that show reduced neutralization by convalescent and post-vaccination sera [3].

Although *in vitro* studies have shown that some variants of concern are less effectively neutralized by sera from vaccinated individuals [4], the clinical implications for post-vaccination breakthrough infection remain largely unknown. In this study, we examined SARS-CoV-2 genomes isolated from individuals identified as vaccine breakthrough cases and compared them to the background of SARS-CoV-2 sequences from Washington over the same time interval.

## Methods

This work was approved by the University of Washington (UW) Institutional Review Board. The UW Virology Lab (UWVL) routinely performs SARS-CoV-2 whole genome sequencing of specimens received for clinical testing. In this study, cases were defined as patients who were fully vaccinated against SARS-CoV-2 (> two weeks post-2^nd^ dose of Pfizer or Moderna vaccine) who subsequently tested positive by RT-PCR. The control group included samples collected in WA from the same time period as the case samples that were sequenced at UWVL. SARS-CoV-2 was detected by RT-PCR as previously described using either the emergency use-authorized UW CDC-based laboratory-developed test, Hologic Panther Fusion or Roche Cobas SARS-CoV-2 tests [5]. Sequencing was attempted on all specimens with Ct < 36 using a multiplexed amplicon sequencing panel from Swift Biosciences [6] or Illumina COVIDSeq (Illumina Inc, USA). Consensus sequences were assembled using a custom bioinformatics pipeline described previously (https://github.com/greninger-lab/covid_swift_pipeline,[6]). We excluded SARS-CoV-2 genome sequences that were incomplete (length < 29kbp) or of low quality (>10% Ns). SARS-CoV-2 lineages and clades were assigned using the PANGOLIN (Phylogenetic Assignment of Named Global Outbreak LINeages, https://pangolin.cog-uk.io/) and NextClade (https://clades.nextstrain.org/) tools. Amino acid changes were annotated according to NextClade. Analysis and calculations were performed in R. Significant differences in case distributions according to variants of concern were determined using Fisher’s exact test.

## Results

Beginning in February 2021, SARS-CoV-2 genome sequencing was requested at UW Medicine hospitals and partners as part of investigations into post-vaccine breakthrough infections. A total of 20 cases were sequenced in this study, including 13 females, 6 males, and 1 with unknown sex. Ages ranged from 26 to 65 years (median=43, IQR=28-58) (Supplementary Table 1), and Ct values ranged from 16.00 to 35.86 (median=20.2, IQR=17.1-23.3). These specimens were collected between February 23 and April 27, 2021. The control group consisted of genomes sequenced by UWVL (n=5174) during the same period as the vaccine breakthrough cohort (Supplementary Table 1).

All 20 of 20 vaccine breakthrough cases were classified as variants of concern: 8 (40%) B.1.1.7, 1 (5%) B.1.351, 2 (10%) B.1.427, 8 (40%) B.1.429, and 1 (5%) P.1 (Figure 1A, Supplementary Table 2). In contrast, during the same time interval, 68% of WA cases sequenced at UWVL represented VOC with 31% B.1.1.7, 1% B.1.351, 3% B.1.427, 27% were B.1.429, and 7% were P.1 (Figure 1B). Overall, variants of concern were proportionally over-represented in breakthrough cases, with the frequency of all VOCs in breakthrough increased 1.47-fold compared to the control group (95% CI [1.45, 1.50], p=0.001). Variants B.1.427, B.1.429, and B.1.1.7 were 3.38-fold (95% CI [0.90, 12.71], p= 0.119), 1.51-fold (95% CI [0.88, 2.59], p=0.203), and 1.29-fold (95% CI [0.75, 2.20], p=0.468) more common in breakthrough cases compared to controls (Supplementary Figure 1, Supplementary Table 2). The overall distribution of variants was significantly different between breakthrough and control groups (p = 0.001, Fisher’s Exact test, Supplementary Table 2). Variants that have been reported to be less effectively neutralized by antibodies from post-vaccinated individuals (P.1, B.1.351, B.1.427, and B.1.429) were identified in 60% of breakthrough cases and 36.7% of control cases, a 1.63-fold change (95% CI [1.14, 2.34], p=0.037).

**Figure 1:**
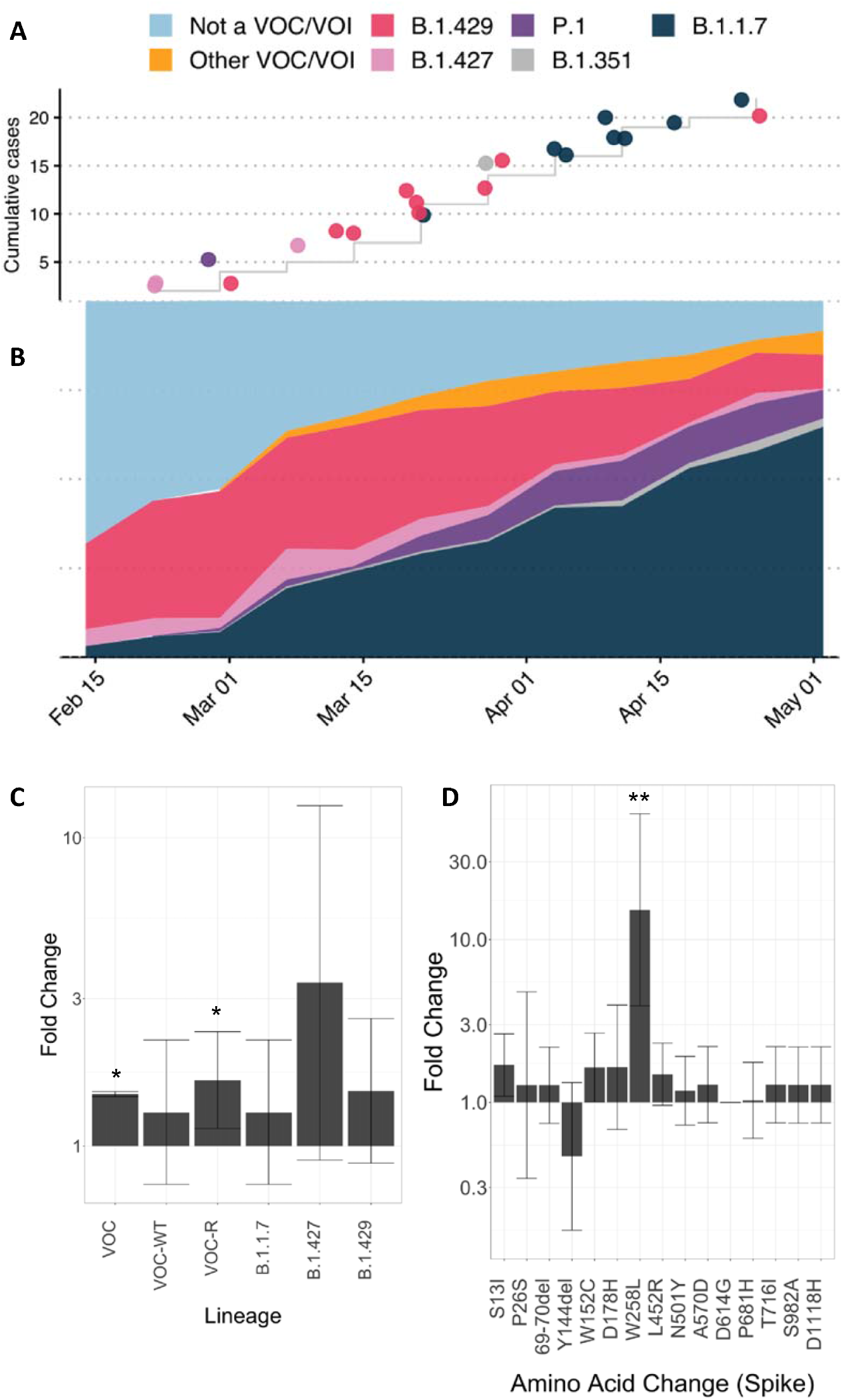
Variants of concern are overrepresented in vaccine breakthrough SARS-CoV-2 genomes. A) Cumulative number of breakthrough cases and their Pangolin lineages. B) Proportions of lineages for UWVL sequences collected during the same time period. Relative risk of vaccine escape for each VOC (C) and spike amino acid change (D). Error bars represent 95% confidence intervals. *p<0.05, **p<0.01 (Fisher’s Exact Test). VOC: variant of concern (total), VOC-WT: variant of concern without reduced neutralization, VOC-R: variant of concern with reduced neutralization

We compared the occurrence of individual mutations in spike in the breakthrough vs. control groups. A single mutation, W258L, was 15.22-fold enriched in breakthrough cases, 95% CI [3.91, 59.10], p=0.008 (Fig. 1D), but was present in only 2 cases. No other mutations showed significant enrichment in the breakthrough group.

Clinical data, including comorbidities and vaccination type and dates (n=19), were available for a subset of subjects (Supplementary Table 3). All patients received mRNA-based vaccines (14 BNT162b2, 5 mRNA-1273); 15 out of 18 reported symptoms, none required hospitalization. Specimens were collected at an average of 67.7 days after vaccination (range=39-112, sd=18.1).

## Discussion

In this study, SARS-CoV-2 variants of concern were found to be overrepresented in vaccine breakthrough cases when compared to cases circulating in the general population of Washington State over the same time interval. Although no single VOC was significantly enriched in the breakthrough cases, subgroup analysis revealed that variants that have shown reduced antibody neutralization *in vitro* (B.1.351, B.1.427, B.1.429, and P1) were overrepresented compared to the B.1.1.7 VOC lineage which is not associated with reduced neutralization. The 20 vaccine breakthrough cases described here also had a substantially stronger viral load than 22 breakthrough cases among Chicago nursing facility staff and residents recently reported by the CDC [7].

The B.1.427/B.1.429 variants were first identified in Los Angeles County, California in July of 2020 [8] and quickly became the dominant form of the virus circulating in that state. These variants were first seen in Washington State in December 2020 and comprised 30% of cases during the study period. The B.1.427/B.1.429 variants are characterized by key spike protein changes, with L452R in receptor binding domain and S13C and W152C in the N-terminal domain (NTD). Recent studies suggest that neutralizing antibody titers from plasma collected from vaccinated or convalescent individuals were reduced 2-7 fold against B.1.427/B.1.429 variant relative to wildtype [8,9]. Similarly, P.1 variant identified in Brazil and B.1.351 variant identified in South Africa also show reduced antibody neutralization efficiency, 4-5 and 5-40 fold, respectively [10,11]. Both P.1 and B.1.351 were uncommon in the control group, with P.1 comprising 6% of cases and B.1.351 less than 1% of cases. In contrast, the B.1.1.7 variant, first identified in the United Kingdom and known to increase infectivity by approximately 50%, is neutralized as efficiently as wild type or only minimally decreased in most studies [11–13]. B.1.1.7 was the major variant circulating in our area during the study period. While it is noted that the introduction of E484K into the B.1.1.7 background has been shown to introduce reduced neutralization efficiency [14], this mutation was not present in any of our breakthrough B.1.1.7 cases, so we considered this strain as having similar neutralization efficiency as wild-type. These data therefore suggest that *in vitro* neutralization assays may predict variants more likely to escape antibodies from vaccinated individuals.

Most mutations thought to lead to antibody neutralization resistance occur in the receptor binding domain of the spike protein [15]. Of note, 2 of 2 B.1.427 cases identified in the breakthrough group contained a spike mutation W258L outside of the receptor binding domain, compared to 50 of 208 (24.03%) in the control group. W258L resides in the so called “NTD-antigen supersite”. While antibodies that bind the receptor binding domain may target different epitopes, a single site of vulnerability to antibody neutralization exists in the NTD [16]. The impact of NTD alterations in antibody neutralization efficiency is largely unstudied. Our data suggest that *in vitro* studies of antibody neutralization residence should include NTD alterations such as W258L.

This study is limited by the small number of subjects in the vaccine breakthrough group, and we cannot exclude the presence of additional breakthrough cases in the control groups due to a lack of clinical data. Moreover, our breakthrough cohort was mostly composed of healthcare workers who were identified by infection control or employee health. In contrast, sequences from UWVL were sampled from across the state and include samples from various testing sites including hospitals, outpatient clinics, and community testing sites.

All 20 breakthrough cases in this study were assigned a lineage without conflicts. This contrasts with a recent study which demonstrated a novel variant in a post-vaccination individual that fell between clades 20B and 20C, raising concern that viral evolution may drive neutralization antibody resistance [2]. Continued surveillance of post-vaccine breakthrough cases may help target variants of concern for inclusion in new or booster SARS-CoV-2 vaccines.

## Data Availability

All sequences have been deposited to GISAID.

## Acknowledgements

Computational analyses were made possible by Scientific Computing Infrastructure at Fred Hutch and UW Laboratory Medicine IT cloud infrastructure.

## Funding

This work is supported by the National Institutes of Health [ORIP grant S10OD028685 to Scientific Computing at Fred Hutch, and UW-Fred Hutch CFAR AI027757 to P.R.].

## Conflicts of Interest

A.L.G. reports contract testing from Abbott Laboratories and research support from Gilead and Merck, outside of the described work.

---

**Supplementary Figure 1.**
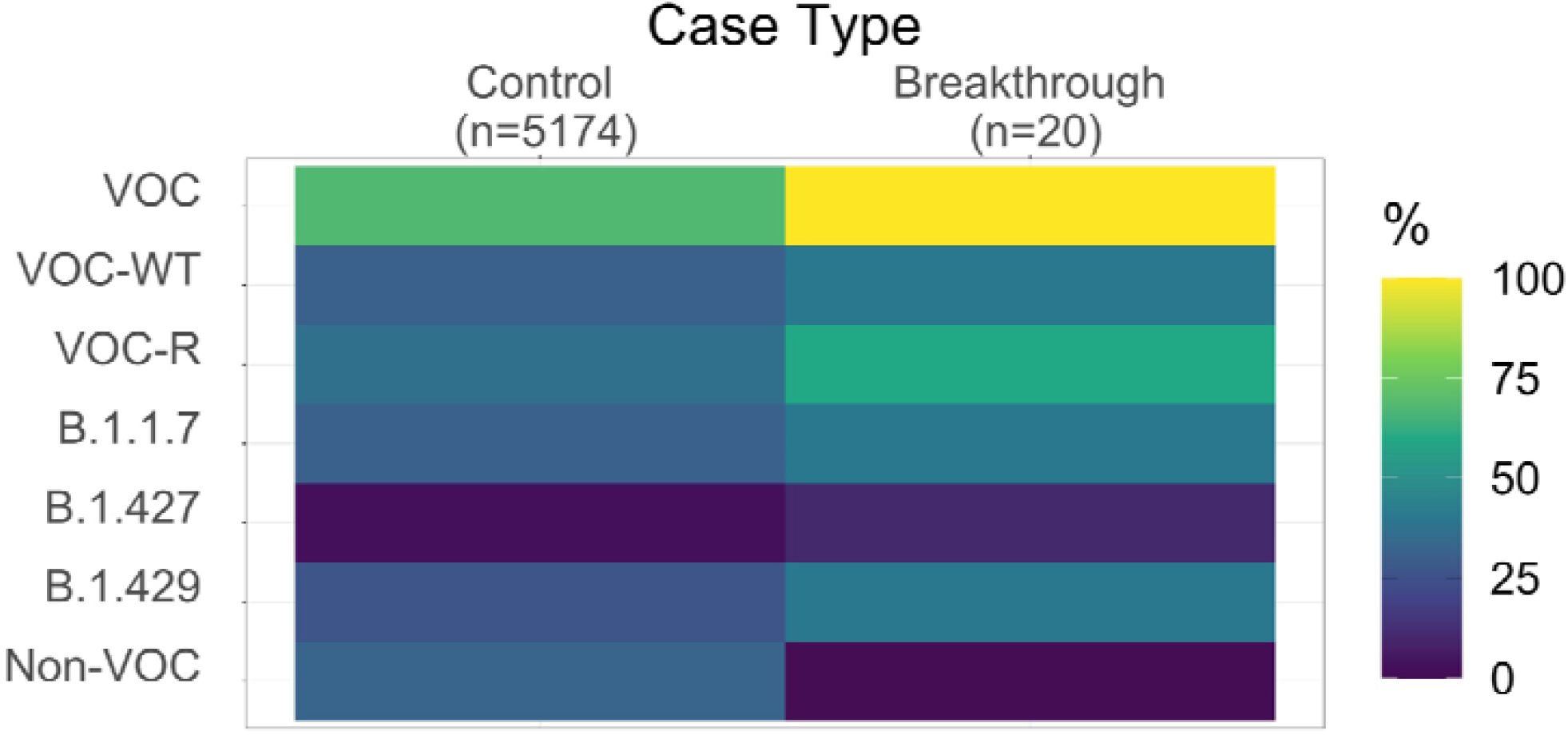
Heat map showing relative frequencies of variants of concern breakthrough vs. control cases.

**Supplementary Table 1:** Study groups

|  | **Vaccine Breakthrough**<br>(n=20) | **UWVL**<br>(n=5174) |
| --- | --- | --- |
| **Age (years)** |  |  |
| Range | 26 - 65 | 0 - 98 |
| Median | 43 | 30 |
| Mean | 43.80 | 32.572 |
| SD | 11.61 | 16.582 |
| IQR | 29.75 | 14.000 |
| Q1 | 34.00 | 21.00 |
| Q2 | 43.00 | 30.00 |
| Q3 | 48.00 | 44.00 |
| Q4 | 65.00 | 98.00 |
| **Sex** |  |  |
| Female | 13 (65.00%) | 2439 (47.14%) |
| Male | 6 (30.00%) | 2592 (50.10%) |
| Unknown | 1 (5.00%) | 143 (2.76%) |
| **Collection date** |  |  |
| Range | 2/23/21 - 4/27/21 | 2/23/21 - 4/27/21 |
| Median | 3/29/2021 | 4/1/2021 |
| Mean | 3/29/2021 | 3/28/2021 |
| IQR | 26.50 | 24.00 |
| Q1 | 3/17/2021 | 3/17/2021 |
| Q2 | 3/29/2021 | 4/1/2021 |
| Q3 | 4/13/2021 | 4/10/2021 |
| Q4 | 4/27/2021 | 4/20/2021 |
| **Ct value** |  |  |
| Range | 16.00 - 35.86 |  |
| Median | 20.16 |  |
| Mean | 21.59 |  |
| SD | 5.32 |  |
| IQR | 6.19 |  |
| Q1 | 17.82 |  |
| Q2 | 20.16 |  |
| Q3 | 24.00 |  |
| Q4 | 35.86 |  |

SD = standard deviation, IQR = interquartile range, Q1= first quartile, Q2 = second quartile, Q3 = third quartile, Q4 = fourth quartile, Ct = PCR cycle threshold

**Supplementary Table 2:** Variants of concern in case vs. control groups

| Vaccine Breakthrough<br>(n=20) |  |  | UWVL<br>(n=5174) |  |  |  |  |
| --- | --- | --- | --- | --- | --- | --- | --- |
| N | 20 | % | N | % | Fold Increase | 95% CI | P* |
| B.1.1.7 | 8 | 40.00% | 1609 | 31.10% | 1.29 | [0.75, 2.20] | 0.001 |
| B.1.351 | 1 | 5.00% | 37 | 0.72% | 6.99 | [1.01, 48.52] |  |
| B.1.427 | 2 | 10.00% | 153 | 2.96% | 3.38 | [0.90, 12.71] |  |
| B.1.429 | 8 | 40.00% | 1372 | 26.52% | 1.51 | [0.88, 2.59] |  |
| P.1 | 1 | 5.00% | 337 | 6.51% | 0.77 | [0.11, 5.20] |  |
| Non-VOC | 0 | 0.00% | 1666 | 32.20% | -- | -- | -- |
| Total-VOC | 20 | 100.00% | 3508 | 67.80% | 1.47 | [1.45, 1.50] | 0.001 |
| VOC-reduced affinity | 12 | 60.00% | 1899 | 36.70% | 1.63 | [1.140, 2.34] | 0.037 |
| VOC-not reduced affinity | 8 | 40.00% | 1609 | 31.10% | 1.29 | [0.750, 2.20] | 0.468 |

236 \*Fisher's Exact Test, VOC = variant of concern

**Supplementary Table 3:**
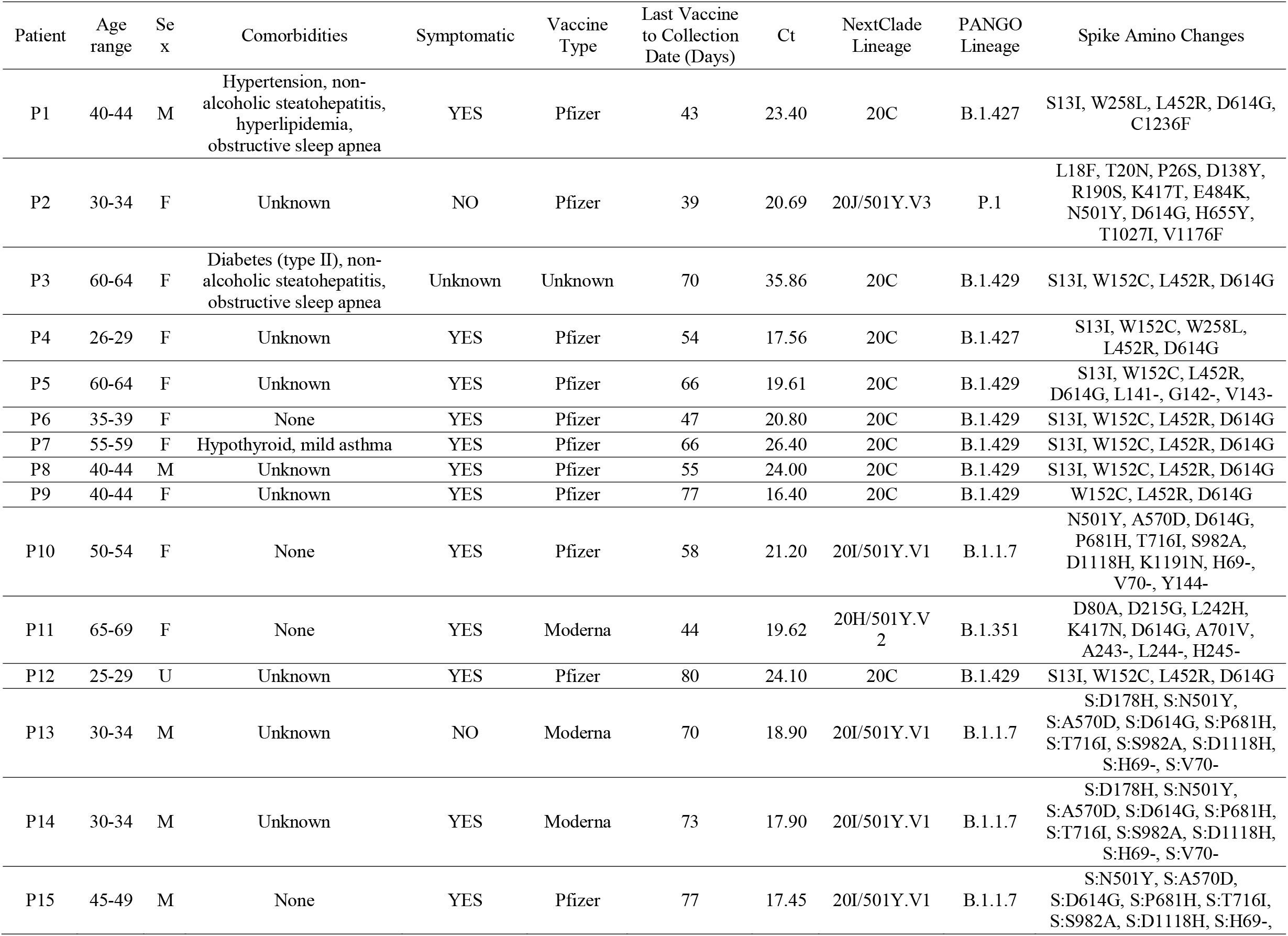
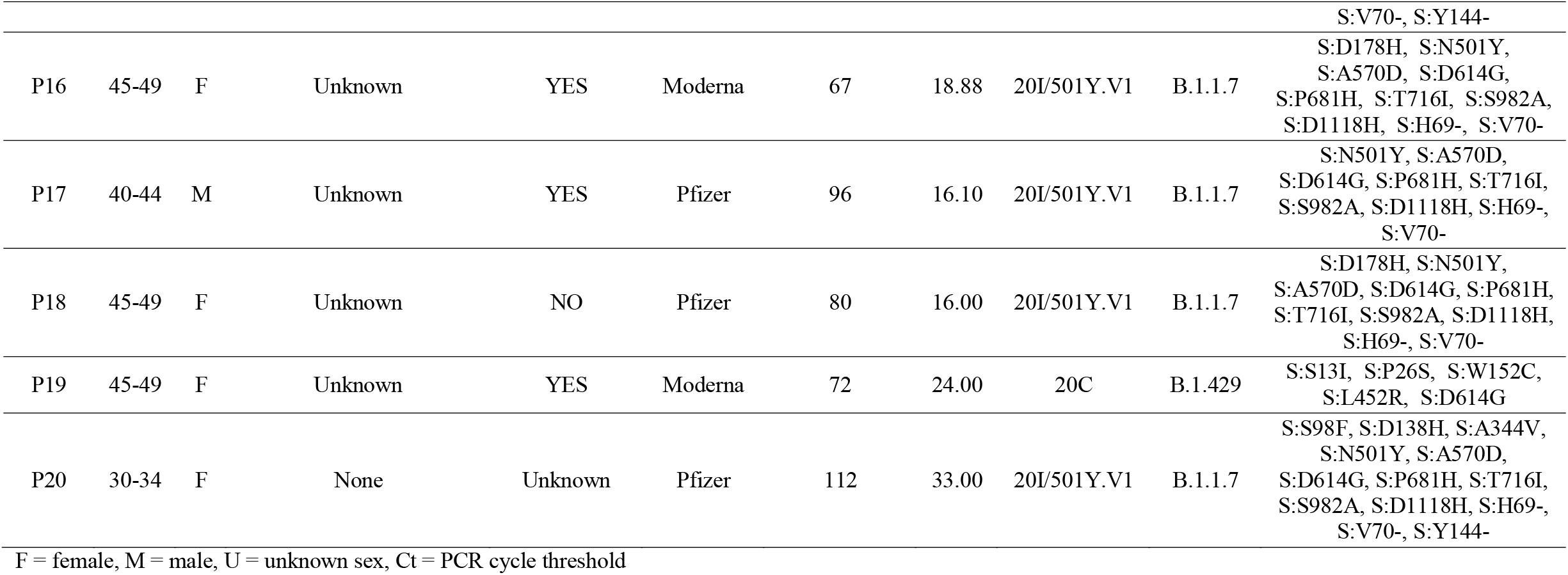
Breakthrough Cases

## Notes

### Author Declarations

This work was approved by the University of Washington Institutional Review Board.

